# Designing and validating a Markov model for hospital-based addiction consult service impact on 12-month drug and non-drug related mortality

**DOI:** 10.1101/2020.12.01.20242164

**Authors:** Caroline A. King, Honora Englander, P. Todd Korthuis, Joshua A. Barocas, K. John McConnell, Cynthia D. Morris, Ryan Cook

**Author notes:** Corresponding Author: Caroline King, MPH, 3181 Sam Jackson Park Road, Portland, Oregon 97209.

## Abstract

**Introduction:** Addiction consult services (ACS) engage hospitalized patients with opioid use disorder (OUD) in care and help meet their goals for substance use treatment. Little is known about how ACS affect mortality for patients with OUD. The objective of this study was to design and validate a model that estimates the impact of ACS care on 12-month mortality among hospitalized patients with OUD.

**Methods:** We developed a Markov model of referral to an ACS, post-discharge engagement in SUD care, and 12-month drug-related and non-drug related mortality among hospitalized patients with OUD. We populated our model using Oregon Medicaid data and validated it using international modeling standards.

**Results:** There were 6,654 patients with OUD hospitalized from April 2015 through December 2017. There were 114 (1.7%) drug-related deaths and 408 (6.1%) non-drug related deaths at 12 months. Bayesian logistic regression models estimated four percent (4%, 95% CI= 2%, 6%) of patients were referred to an ACS. Of those, 47% (95% CI= 37%, 57%) engaged in post-discharge OUD care, versus 20% not referred to an ACS (95% CI= 16%, 24%). The risk of drug-related death at 12 months among patients in post-discharge OUD care was 3% (95% CI= 0%, 7%) versus 6% not in care (95% CI = 2%, 10%). The risk of non-drug related death was 7% (95% CI =1%, 13%) among patients in post-discharge OUD treatment, versus 9% not in care (95% CI= 5%, 13%).

**Discussion:** Our novel Markov model reflects trajectories of care and survival for patients hospitalized with OUD. This model can be used to evaluate the impact of other clinical and policy changes to improve patient survival.

## Introduction

Drug overdose is the leading cause of unintentional injury death in the United States (1). Among people with opioid use disorder (OUD), 20% eventually die of drug overdose (2), but cardiovascular diseases, cancer, and infectious diseases also contribute. Patients with OUD who are hospitalized for OUD-related and other diagnoses are often medically complex and face life-threatening illnesses. These patients experience higher mortality rates than hospitalized patients with similar conditions (2).

Hospitalization is a vulnerable time for patients with OUD. People with OUD may leave the hospital before completing recommended medical therapy if withdrawal symptoms are untreated (3). People who withdraw from opioids have lower drug tolerance and increased risk of drug overdose after discharge in the absence of treatment for OUD (4-6). Medications for opioid use disorder (MOUD) delivered in the hospital can treat withdrawal symptoms and reduce overdose risk (7), and are often necessary, but not sufficient, to help keep patients engaged in inpatient care. Despite this, most hospitalized patients with OUD are not started on MOUD (8, 9), though, when offered, nearly three-quarters of patients with OUD choose to start MOUD (10). Interventions to improve initiation of MOUD among hospitalized patients are urgently needed (11).

Addiction consult services (ACS) are an emerging intervention to engage hospitalized patients in care and meet patient-driven goals for substance use treatment (12). Evaluation of ACS demonstrates improved engagement in post-hospitalization treatment and decreased substance use (11, 12). However, assessing the effect of ACS using gold-standard study designs is challenging because of the costs and logistical challenges associated with multi-site, cluster-randomized trials. Additionally, it can be difficult statistically to assess distal, rare outcomes like drug-related mortality in the context of a hospital-based intervention. We consequently do not know how ACS affect post-discharge drug-related mortality or non-drug related mortality for patients with OUD.

Modeling allows researchers to rapidly test different care delivery scenarios and capture robust estimates of study outcomes, which can support healthcare system decision-making and answer salient clinical questions in the midst of the opioid overdose epidemic. Modeling inpatient care scenarios can guide healthcare systems in addressing a rapidly evolving epidemic more quickly and adaptively than randomized trials. Simulation modeling has previously been used to estimate prevented overdose deaths from the expansion of naloxone distribution (13-15), and the implementation of safe-injection sites (16). The objective of this study was to design and validate a Markov model that estimates the impact of ACS care on 12-month mortality among hospitalized patients with OUD.

## Methods

### Model Structure

We used a Markov model to estimate the impact of ACS care on 12-month mortality among hospitalized patients with OUD (Figure 1). Our model has the following components: ACS consult, post-discharge OUD treatment engagement, and 12-month post-discharge drug related death, non-drug related death, and survival.

**Figure 1.**
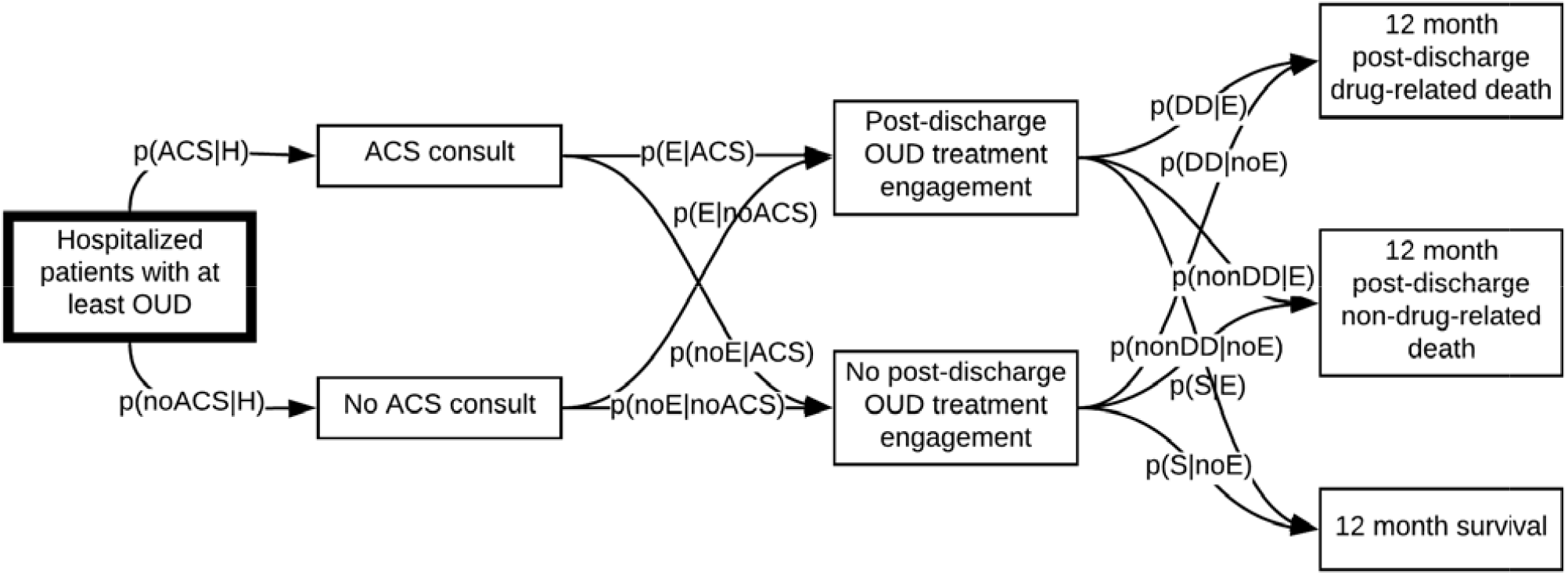
Markov model of hospital-based addiction care in Oregon, 2015-2018.

The Oregon Health & Science University’s Institutional Review Board approved this study (#00010846).

#### ACS Referral

Once patients are admitted to the hospital, they can be referred to ACS care. ACSs exist across a growing number of North American hospitals. Typically, they include care from an interprofessional team that may include of medical providers, social workers, nurses, and alcohol and drug counselors (17). Some intentionally include people with lived experience in recovery (18-20). ACSs typically address needs of people use any substance (for example, stimulant, alcohol, and opioids) and care includes comprehensive assessments, withdrawal management, medication treatment, psychosocial and harm reduction interventions, and efforts to support patient engagement and linkage to care across settings. ACSs commonly also provide staff education and patient advocacy (17, 21, 22). For this model, we used an intention-to-treat approach; all patients referred to ACS were included regardless of level of care engagement or specific services received.

#### Post-discharge OUD treatment engagement

We used a modified Healthcare Effectiveness Data and Information Set (HEDIS) measure of engagement to stratify for post-discharge OUD treatment engagement. The original measure requires that patients initiate treatment and have two or more additional alcohol or drug services or medication for OUD within 34 days of initiation (23). Recent research has shown that evidence-based MOUD has superior outcomes in preventing mortality and decreasing opioid use (7). For this reason, we defined post-discharge OUD treatment engagement as: 1) at least two filled prescriptions for buprenorphine, extended-release naltrexone, or methadone from an Opioid Treatment Program in the 30 days following hospital discharge, or 2) a prescription for extended-release naltrexone or buprenorphine that covered 28 of the 30 days post-hospital discharge (24).

#### 12-month mortality

At twelve months, deaths are classified as drug related versus non-drug related (including as circulatory, neoplasm, infectious, digestive (including alcohol-related liver disease), external (including suicide and unintentional injury), respiratory, endocrine, and other) by ICD-10 mortality codes described by Hser et al (2).

### Model data

Where data exists for recalibration, our Markov model could be used in any setting with patients hospitalized with OUD. We populated our model with data from Oregon Medicaid claims data and expert opinion, described below, to reflect care from an addiction consult service in Portland, Oregon, and its impact on post-discharge drug and non-drug related mortality.

### Setting and study design

Oregon Health & Science University in Portland, Oregon is home to an inpatient ACS, the Improving Addiction Care Team (IMPACT). IMPACT is a hospital-based service that utilizes an interdisciplinary team of physicians, advanced practice providers, social workers, and peers with lived experience in recovery to support non-treatment seeking adults with substance use disorder. Patients are eligible to be referred if they have known or suspected substance use disorder (SUD), other than tobacco use disorder alone. IMPACT conducts substance use assessments, initiates medication-based treatment (including buprenorphine, methadone and extended release naltrexone for OUD) and behavioral treatment where appropriate, and connects patients to post-discharge SUD treatment. IMPACT utilizes a harm reduction approach and integrates principles of trauma-informed care. Previous research describes IMPACT’s design and evaluation (10, 11, 19-21, 25, 26). Notably, IMPACT is the only comprehensive ACS in Oregon, though a few hospitals offer MOUD initiation during hospitalization.

### Participants

We used Oregon Medicaid claims data to identify patients hospitalized at least once with OUD from April 2015 through August 2018, including IMPACT patients. For mortality analyses only, we also utilized mortality data from Oregon Vital Statistics through December 31, 2018; thus, patients admitted through January 1, 2018 were included to allow 12 months of follow-up time. Patients were eligible for inclusion if they were over 18 years old and had an ICD-9 (304.*) or ICD-10 (F11*) diagnosis of OUD during a hospital admission.

#### Cohorts for transition points

We defined three cohorts for our analyses utilizing Oregon Medicaid data. First, we included all patients who met eligibility criteria in analysis for our first transition, referral to ACS. Then, we used a matched cohort of three controls to one IMPACT patient for our post-discharge OUD care engagement and mortality analyses. We matched without replacement on hospital admission quarter and admission number, including one admission per person.

### Transition data

For ACS referral, we identified all hospitalized patients with OUD in Oregon during the study period, and then identified the subset who were referred to the ACS. For post-discharge OUD treatment engagement, we used Oregon Medicaid claims data to identify if patients met the modified HEDIS engagement measure in the 30 days following hospital discharge. For 12-month mortality, we used Oregon Vital Statistics data to identify deaths in our cohort during the study period through December 31, 2018. For mortality models, the cohort was limited to include only participants seen before January 1, 2018 to allow for 12 months of follow-up time for all participants. We classified deaths as drug related versus non-drug related as indicated above. We manually reviewed deaths that were not captured by these codes and reclassified to fit into drug versus non-drug related categories.

### Transition probabilities

We used a Bayesian approach to obtain transition probabilities for our Markov model using Oregon data. First, we obtained prior information from experts in addiction (described below). After surveying expert participants, we calculated the mean and identified the minimum and maximum ratings. We then numerically fit beta distributions to those quantities using differing “confidence levels” (27). Then, we updated our priors with the information from data about our cohort described above. We estimated marginal probabilities over observed cases using fitted Bayesian logistic regression models at each transition point (28).

#### Bayesian priors via expert elicitation

We used expert elicitation to capture prior information for our models. We identified important covariates at each transition point, including age (in years), gender (female/male), race (White/not White/unknown), ethnicity (Hispanic/Not Hispanic), concurrent alcohol use disorder (yes/no), concurrent stimulant use disorder (yes/no), hospital length of stay (in days), rural residence (yes/no), filled at least one prescription for medication for OUD in the month before hospital admission (yes/no), previously admitted to the hospital (yes/no), and Chronic Illness and Disability Payment System (CDPS) Score (continuous). The engagement model also included referral to an ACS (yes/no). The mortality models included engagement in care after discharge (yes/no) and filled a naloxone prescription in the 30 days after hospital discharge (yes/no).

We used a clinical-vignette design to ask providers about the relevance of covariates on patient outcomes. To do this, participants provided a probability estimate for different events: referral to an ACS, post-discharge engagement, and mortality.

For example, a vignette could read:

> *“The patient is a young White man with OUD and AUD. He was in the hospital for several days. He was on medication for OUD at admission. He had never previously been admitted to the hospital. He has many comorbidities. He is not from a rural area. What is the probability he engaged in post-discharge treatment for OUD within 30 days of discharge?”*

Experts evaluated 16 (referral to ACS), 17 (engagement) and 18 (mortality) vignettes selected from an optimal experimental design generated for each model (29). From the optimal design, we chose a subset of the vignettes that were substantially different from one other for ease of interpretability and to maximize the information gathered about each covariate.

As part of our IRB-approved research, study authors (HE, PTK) generated lists of experts in addiction consult services and hospital-based addiction treatment in general in the United States. Each participant took only one survey. We aimed to recruit at least five participants for each survey, with a goal of at least three responses per survey. For the referral to ACS survey, we also asked participants to refer hospitalists at their institutions to complete the survey, as hospitalists are frequently providers who refer patients to ACS. Ultimately, six participants took the ACS survey (6 of 11, 54.5%), four took the engagement survey (4 of 5, 80%), and three took the mortality survey (3 of 8, 37.5%).

#### Bayesian logistic regression models

We used the transformed prior information from expert surveys and Oregon Medicaid cohort data to fit Bayesian logistic regression models at each transition point. Models were fit using Markov Chain Monte Carlo methods (30). We sampled each parameter 10,000 times with 2000 burn-in chains. We used multiple metrics to assess model convergence. First, we used Gelman and Rubin’s potential scale reduction factor; all values in all models equal 1.0. Values close to 1.0 are suggestive of convergence. Effective sample sizes all approximated the number of posterior draws requested. All model trace plots appear to have a caterpillar-like distribution, and there were no divergent transitions. Autocorrelation plots for all parameters suggest low autocorrelation. We used the package Shiny Stan to evaluate Bayesian model fit (31).

We tested different prior information strengths: first, using a cohort sample size method, where the prior information equivalates a percent of the study sample size (0.1%, 1%, 5% and 10%); second, using a confidence interval method, where we fit beta distributions to the range of survey responses, and then used the maximum and minimum values as borders for 80%, 85%, 90%, and 95% confidence intervals. We picked the best-fit model using Pareto smoothed importance-sampling leave-one-out cross validation using the loo package in R where lower expected log predictive density values indicate a better model fit (32). We also prioritized models where Pareto k diagnostic values had at least good reliability for all estimates.

We used mcmcObsProb in the BayesPostEst package (33) to estimate marginal transition probabilities over observed cases with the fitted Bayesian logistic regression models. We created prior-posterior plots using ggplot2 (34).

### Model validation

We validated our model using the frameworks suggested by the International Society for Pharmacoeconomics and Outcomes Research and the Society for Medical Decision Making’s Good Research Practices Model Validation guidelines (ISPOR-SMDM) (35). We explored five components of validity: face validity, internal validity, cross validity, predictive validity, and external validity. As suggested, we provide a non-technical description of our model in Appendix 3.

## Results

There were 8,450 patients admitted at least once with OUD in Oregon from April 2015 through August 2018. Among the 6,654 patients seen by January 1st, 2018, at twelve months, 114 (1.7%) participants died from drug-related causes and 408 (6.1%) died from non-drug related causes. Participant demographics are included in Table 1.

**Table 1.** Participant demographics.

|  | **All patients**<br>n=8,450 | **Seen by ACS**<br>n=265 | **Not Seen by ACS**<br>n=8,185 | **p-value** |
| --- | --- | --- | --- | --- |
| **Age Years** | 44.5 (15.4) | 39.5 (0.77) | 44.6 (0.17) | <0.001 |
| **Gender Male** | 3,632 (43.0%) | 159 (60.0%) | 3,473 (42.4%) | <0.001 |
| **Race White** | 5,919 (70.1%) | 169 (63.8%) | 5,750 (70.3%) | 0.034 |
| Not White | 543 (6.4%) | 16 (6.0%) | 527 (6.4%) |  |
| Unknown race | 1,988 (23.5%) | 80 (30.2%) | 1,908 (23.3%) |  |
| **Ethnicity**<br>Hispanic | 299 (3.5%) | 10 (3.8%) | 289 (3.5%) | 0.002 |
| **Alcohol use disorder** | 306 (3.6%) | 14 (5.3%) | 322 (3.9%) | 0.269 |
| **Stimulant use disorder** | 689 (8.2%) | 41 (15.5%) | 642 (7.8%) | <0.001 |
| **Length of stay (days)** | 6.6 (11.2) | 14.9 (0.97) | 6.4 (0.12) | <0.001 |
| **Rural residence** | 2,234 (26.4%) | 32 (12.1%) | 2,202 (26.9%) | <0.001 |
| **Medication for OUD at hospital admission** | 1,508 (17.8%) | 48 (18.1%) | 1,460 (17.8%) | 0.908 |
| **Previously admitted to hospital** | 1,891(22.4%) | 116 (43.8%) | 1,775 (21.7%) | <0.001 |
| **CDPS Score** | 2.5 (1.6) | 3.11 (0.11) | 2.48 (0.02) | <0.001 |

Transition probabilities derived from Bayesian logistic regression models are depicted in Figure 2. In our study, 4% (95% CI= 2%, 6%) of patients admitted at least once for OUD were referred to an ACS in Oregon. Of those, 47% (95% CI= 37%, 57%) engaged in post-discharge OUD care. Of the 96% not seen by an ACS, 20% (95% CI= 16%, 24%) engaged in post-discharge OUD care. The risk of drug-related death at 12 months among patients who engaged in post-discharge OUD care was 3% (95% CI= 0%, 7%) versus 6% (95% CI = 2%, 10%) in patients who did not engage in care. The risk of non-drug related death was 7% (95% CI =1%, 13%) among patients who engaged in OUD treatment, versus 9% (95% CI= 5%, 13%) for those who did not. For referral to ACS care, the best-fit Bayesian logistic regression model used an 80% confidence interval; for all other models, a sample size of 0.1% fit best (Appendix 1). All estimates had acceptable Pareto k-diagnostic values. We report posterior intervals for each covariate from Bayesian logistic regression models in Appendix 2.

**Figure 2.**
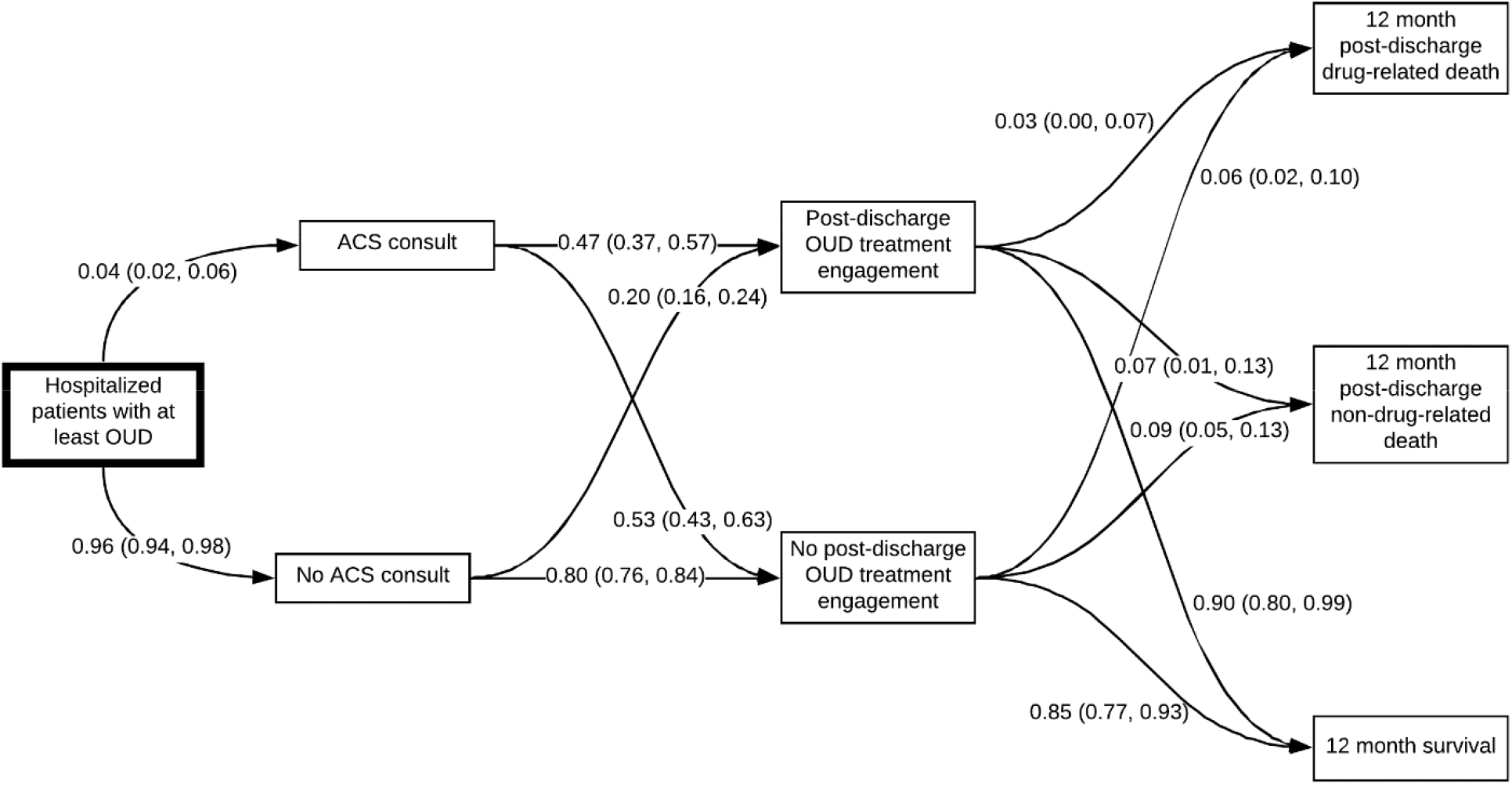
Markov model with estimated transition probabilities for hospital-based addiction care in Oregon, 2015-2018.

### Model validation

#### Face validity

To assess face validity, one researcher (CK) designed the model and received feedback from experts in addiction medicine outside of the study team about the model’s face validity. Experts agreed that the model reflected the path of care for patients admitted to hospitals in Oregon with OUD *(structure)*. Further, the use of Oregon Medicaid data, versus data from the literature, was considered a strength in deriving *evidence* for the model by outside experts. ACS and their impact on care for patients with OUD is of immense interest to healthcare systems and policymakers, and experts also agreed that the question was timely and important *(problem formation)*. Finally, after data analysis, the model results were presented to researchers who agreed that estimates from the model matched their expectations (results).

#### Internal validity

We conducted additional checks and analyses to ensure internal validity of our Bayesian approach (also referred to as technical validity, (36)). First, a recent paper used a similar approach and data structure to evaluate the impact of prenatal maternal factors on nonadherence to infant HIV medication in South Africa. After building our Bayesian model, we used the deidentified data from the South Africa analysis to attempt to replicate identical results as were published. The built model exactly replicated the results of the South African analysis. Second, we conducted classic logistic regression models for each transition point in addition to the Bayesian models. We placed a 1/3, 1/3 noninformative prior (Kerman’s prior) on all covariates, which should be roughly approximate to the classic logistic regression results. Our results with non-informative priors were sufficiently similar to classical logistic regression results. Finally, we conducted code “walk throughs” as suggested, where the analyst (CK) walked through code with an expert in these methods (RC).

In addition to the above steps, because we used Bayesian analyses for our transition probabilities, we needed to ensure that our final estimates of confidence intervals around engagement and mortality estimates actually encompassed the observed number of people who engaged, and people who died from drug-related and non-drug related deaths. Of the 6,654 patients with 12 months follow-up time, the model estimates that 1,330.8 patients engage in care (95% CI =1,064.6, 1,597.0). We observed 1,318 patients who engaged in care in the cohort. Additionally, the model estimated 357.2 drug related deaths (95% CI= (98.5, 632.6)); there were 114 observed drug related deaths in the dataset. Similarly, the model predicted 570.8 non-drug related deaths (95% CI= 263.6, 865.0)); there were 408 observed non-drug related deaths in the dataset. Mortality analyses rarely account for all sources of follow-up which may mean that reported mortality estimates in the literature are lower than in reality. Thus, it was not surprising that modeled transition probabilities from Bayesian logistic regression for 12-month mortality may be higher than raw observed proportions.

#### Cross-validation

Researchers at a separate academic medical center have developed, validated and calibrated the Reducing Infections Related to Drug Use Cost-Effectiveness (REDUCE) model, a Monte Carlo microsimulation model (37). This model has the capacity to answer similar questions to what we post here, using estimates derived from published data and from expert sources. In contrast to our model which uses a cohort defined by opioid use disorder, the REDUCE model simulates data for people who inject drugs. Because model estimates for the REDUCE model are derived from a variety of sources in different parts of the county, we expected outcomes from the REDUCE model to be different from our model; we felt these differences are important to understand.

To support cross-validation of our model, the research team that developed the REDUCE model generated 4,153 simulated patients admitted to the hospital for the first time. Of those, 36 died while in the hospital (0.9%). Of the 4117 still alive at hospital discharge, 96 (2.3%) died within 12 months of hospital discharge (95% CI = 1.9%, 2.8%). This is lower than our estimated 928 (13.9%) deaths from our Markov model (95% CI = 5.4%, 22.5%).

There are several important differences between the REDUCE model and our model. First, as previously mentioned, the REDUCE model simulates data from patients who inject drugs, while ours models patients who have OUD more generally. There are important demographic differences between these two groups, including that our model also includes patients with a primary diagnosis of cancer. Next, the percentage of people seen by an ACS in the REDUCE model was higher than in our model: 25% of patients in REDUCE were seen by an ACS versus 4% in our model. The REDUCE model uses data from Boston, where higher numbers of patients are seen by ACS. This makes it challenging to understand REDUCE estimates in the context of Oregon specifically. Additionally, patients had a higher post-discharge treatment engagement rate in the REDUCE model. In REDUCE, approximately 25.2% of patients receive medication for OUD for at least one week in the month following discharge, versus our model, where 20% of patients not seen by an ACS receive MOUD after discharge. Finally, data from the first simulated admission was used to estimate 12-month mortality from REDUCE; because we matched our cohort controls on the number of previous admissions among patients seen by an ACS, it is possible that our patients were older and sicker than patients who had never previously been admitted to the hospital. While the base model structures are similar, our model is populated with data that provides a focused understanding of addiction consult services in Oregon. Populating our model with different data, including Boston estimates, could provide tailored explorations of ACS in different settings.

#### External validity

To examine external validity, we used large, high-quality, recent studies of representative populations in independent cohorts of participants to separately validate post-discharge OUD treatment engagement and 12-month drug related and non-drug related mortality. We simulated a cohort of size determined from outside research and looked to see if our simulated confidence interval (cohort simulation/matrix multiplication method, (36)) was different from observed values or confidence intervals from the published estimates (Table 2). Where there was disagreement, we describe potential causes.

**Table 2.** Table of results for external validation of Markov model.

| **Data Source** | **Justification of selection** | **Dependent, partially dependent, independent data source** | **Part of model evaluated** | **Comparison of differences and results in data sources** | **Evaluation of cohort simulation results versus observed data** |
| --- | --- | --- | --- | --- | --- |
| Naeger et al. (38) | Testing in national dataset | Independent | Post-discharge OUD treatment engagement | Data from 36,719 patients with an inpatient admission for opioid abuse, dependence, or overdose, 2010 to 2014<br>-Data from time period just prior to Oregon Medicaid cohort; engagement may have been lower<br>-Included any prescription for post-discharge MOUD | Cohort simulation showed 7343.8 (95% CI = 5875, 8812) people predicted to engage versus 6132 people observed<br>-Modeled confidence interval contains point estimate of observed engagement |
| LaRochelle et al 2018 (39) | Testing in large cohort study | Independent | 12-month drug and non-drug related mortality | 17,568 Massachusetts adults without cancer from 2012 to 2014<br>-Dataset mortality may be lower because of exclusion of patients with cancer<br>-Post-discharge treatment engagement for OUD included all time, to 12 months, of post-discharge engagement, which may further decrease drug-related deaths | Cohort simulation showed 8.6 non-drug related deaths per 100 person-years (95% CI=1.5, 13.0), and 5.4 opioid-related deaths per 100 person-years (95% CI=4.0, 9.5)<br>-Observed all-cause mortality was 4.7 deaths (4.4, 5.0) per 100 person-years; opioid-related mortality was 2.1 deaths (1.9 to 2.4) per 100-person years<br>-There is no difference in non-drug related deaths between the simulated cohort and observed data |
|  |  |  |  |  | -Opioid-related deaths may be higher in our model because of a more liberal definition of opioid-related deaths |
| Ashman et al. (CDC) (40) | Testing in large cohort study | Independent | 12-month all-cause mortality | -24,340 patients with an opioid hospitalization across 94 National Hospital Care Survey hospitals<br>-Analysis included patients with cancer | Cohort simulation showed 3,394 all-cause deaths (95% CI = 1324, 5478) versus 1,879 (2,295*0.819) all-cause deaths observed<br>-Modeled confidence interval contains point estimate of observed all-cause mortality |

#### Predictive validity

All relevant data was included in building the Markov model described in this paper. We have planned analyses to evaluate our model predictions versus Medicaid claims data for the same cohort of patients seen in through December of 2020, once data is released.

## Discussion

We built and validated a Markov model that reflects trajectories of care and survival at twelve months for patients hospitalized with OUD in Oregon. We used a Bayesian framework to integrate clinical expertise with data from Oregon Medicaid claims to estimate transition probabilities in our model. After development, we validated our model using ISPOR-SMDM standards, evaluating face validity, internal validity, cross validity, predictive validity and external validity.

The single other model that evaluates ACS care delivery is the REDUCE model, used in model cross validation in this analysis (37). Versus the REDUCE model, our model estimates are more context-relevant estimates of post-discharge OUD treatment engagement and 12-month drug and non-drug related mortality in Oregon. Our overall mortality estimate is higher than the REDUCE model, which may reflect severity of illness of people who are older, sicker, with more previous inpatient hospitalizations and limited linkage to post-discharge OUD care in Oregon. This is important as one potential use of our populated model is to predict the impact of expanding inpatient ACS care in Oregon; a model populated with Oregon data may better reflects the local care setting at baseline may provide more accurate results following intervention. Additionally, populating our model with different data in different ACS context may similarly provide tailored results.

This study had several limitations. First, because we sought to build a model that reflected addiction care in Oregon, the model may not be generalizable to other settings. Still, the Oregon experience may help inform modeling in other states with limited ACS uptake, and we used Bayesian estimates from national experts to inform transition probabilities. Second, claims data is often inaccurate in classifying patient race and ethnicity; our study estimates may not correctly capture the experience of people of color in Oregon. Third, we originally planned to use 30-day mortality as an outcome for this study, but we were unable to do so because of limited drug-related mortality in the 30-day post-discharge period; we used 12-month mortality data instead. Finally, Medicaid claims data does not separate costs for inpatient delivery of medication for OUD, so it was not possible to tell if patients received OUD inpatient outside of an ACS.

This model can be used to evaluate changing scenarios of care in spaces where healthcare providers, healthcare systems, or policymakers are considering implementing or changing ACS coverage in their applicable system. The strength of the model comes from the estimates used to populate it, and with recalibration, the model can be adapted to different settings of ACS care delivery. In this paper, we describe data that reflects ACS care in Oregon. Using this data, we can model changing scenarios of care in Oregon, from increasing ACS care delivery to implementing drug-policy related changes, potentially including reducing barriers to naloxone access, implementing safe consumption sites or safe supply interventions, and others. Future research should use this model to evaluate changes in care delivery in Oregon to understand how these changes may impact survival among patients with OUD.

## Conclusion

Hospitalization is a critical time for patients with OUD, and addiction consult services can help support patients during hospitalization and connect them to post-discharge care. Markov modeling can help researchers, clinical teams and policy makers understand how changes in care systems might impact patient outcomes. Additionally, our model allows healthcare systems and policymakers to evaluate the impact of ACS on mortality. In this work, we built and validated a Markov model that reflects the trajectories of care and survival for patients hospitalized with OUD in Oregon. Future research should use this work to evaluate state-wide clinical and policy changes that may impact patient survival.

## Data Availability

Data may be available pending appropriate data access permissions and approval through OHSU's IRB.

## Acknowledgements

We would like to thank Dr. Christina Nicolaidis, MD, MPH, for her guidance and support for this project.

### Appendix 1. Model fit statistics

|  | Sample Size or Confidence Interval percentage | Expected log pointwise predictive density | Effective number of parameters | Leave-one-out information criterion | Pareto k diagnostic values (all at least "ok") |
| --- | --- | --- | --- | --- | --- |
| Referral to ACS | **0.1% sample size** | -1108.8 | 10.6 | 2217.6 | Yes |
|  | **1% sample size** | -1273.0 | 6.9 | 2546.1 | Yes |
|  | **5% sample size** | -1614.2 | 5.6 | 3228.5 | Yes |
|  | **10% sample size** | -1949.8 | 7.4 | 3899.5 | Yes |
|  | **80% confidence interval** | -1103.8 | 11.2 | 2207.6 | Yes |
|  | **85% confidence interval** | **-1109.9** | **10.7** | **2219.7** | **Yes** |
|  | **90% confidence interval** | -1118.6 | 10.6 | 2237.2 | Yes |
|  | **95% confidence interval** | -1132.2 | 10.4 | 2264.5 | Yes |
| Engagement in post-discharge OUD treatment | **0.1% sample size** | **-397.1** | **12.4** | **794.2** | **Yes** |
|  | **1% sample size** | -406.6 | 10 | 813.3 | Yes |
|  | **5% sample size** | -457.8 | 7.4 | 915.7 | Yes |
|  | **10% sample size** | -511.3 | 7.2 | 1022.5 | Yes |
|  | **80% confidence interval** | -415.6 | 9.6 | 831.3 | Yes |
|  | **85% confidence interval** | -419.2 | 9.5 | 838.5 | Yes |
|  | **90% confidence interval** | -424.3 | 8.9 | 848.6 | Yes |
|  | **95% confidence interval** | -434.0 | 8.4 | 868.0 | Yes |
| **Twelve-month drug-related mortality** | **0.1% sample size** | **-95.9** | **8.7** | **191.8** | **Yes** |
|  | **1% sample size** | -98.5 | 6.3 | 197.1 | No |
|  | **5% sample size** | -109.1 | 5 | 218.2 | No |
|  | **10% sample size** | -116.6 | 5.1 | 233.1 | No |
|  | **80% confidence interval** | -104.3 | 6.6 | 208.7 | No |
|  | **85% confidence interval** | -104.9 | 6.3 | 209.8 | No |
|  | **90% confidence interval** | -106.1 | 6.2 | 212.3 | No |
|  | **95% confidence interval** | -107.3 | 5.8 | 214.6 | No |
| **Twelve-month non-drug-related mortality** | **0.1% sample size** | **-189.4** | **14.0** | **378.8** | **Yes** |
|  | **1% sample size** | -189.9 | 11.4 | 379.8 | Yes |
|  | **5% sample size** | -192.8 | 8.7 | 385.7 | Yes |
|  | **10% sample size** | -194.3 | 7.4 | 388.7 | No |
|  | **80% confidence interval** | -195.2 | 7.6 | 390.4 | Yes |
|  | **85% confidence interval** | -195.5 | 7.2 | 391.0 | Yes |
|  | **90% confidence interval** | -196.1 | 7.0 | 392.2 | Yes |
|  | **95% confidence interval** | -196.9 | 6.7 | 393.8 | No |

### Appendix 2. Estimates from classical and Bayesian logistic regression models, and prior-posterior plots

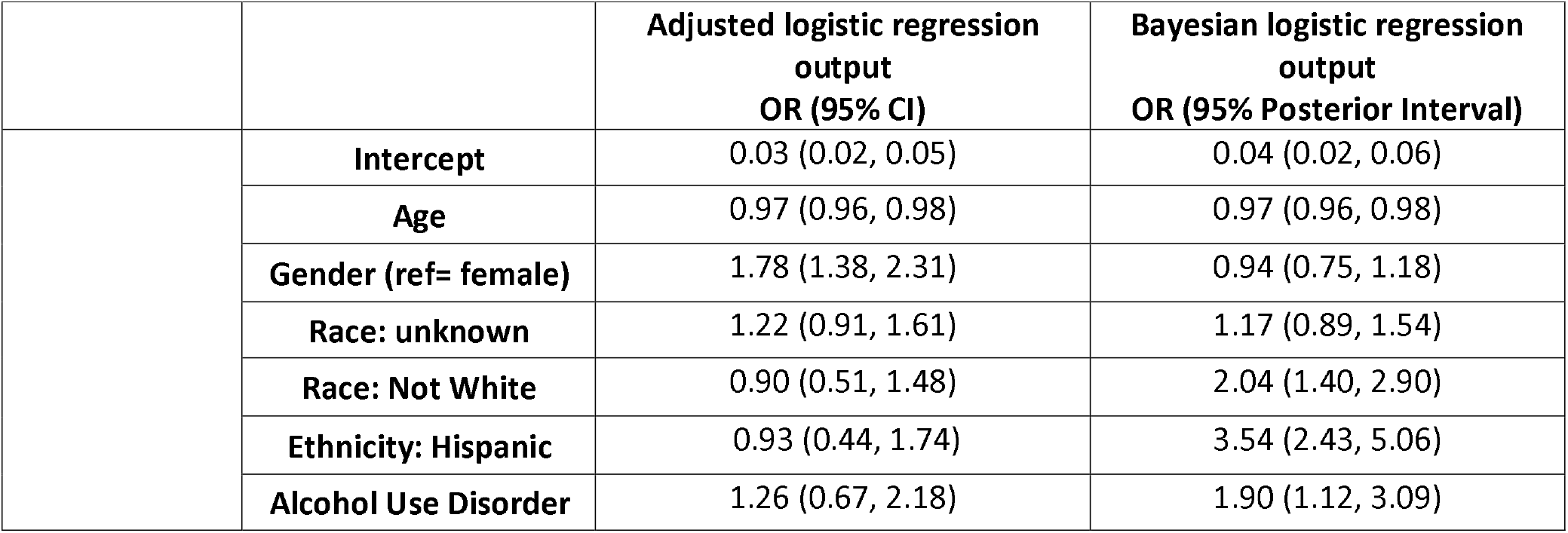
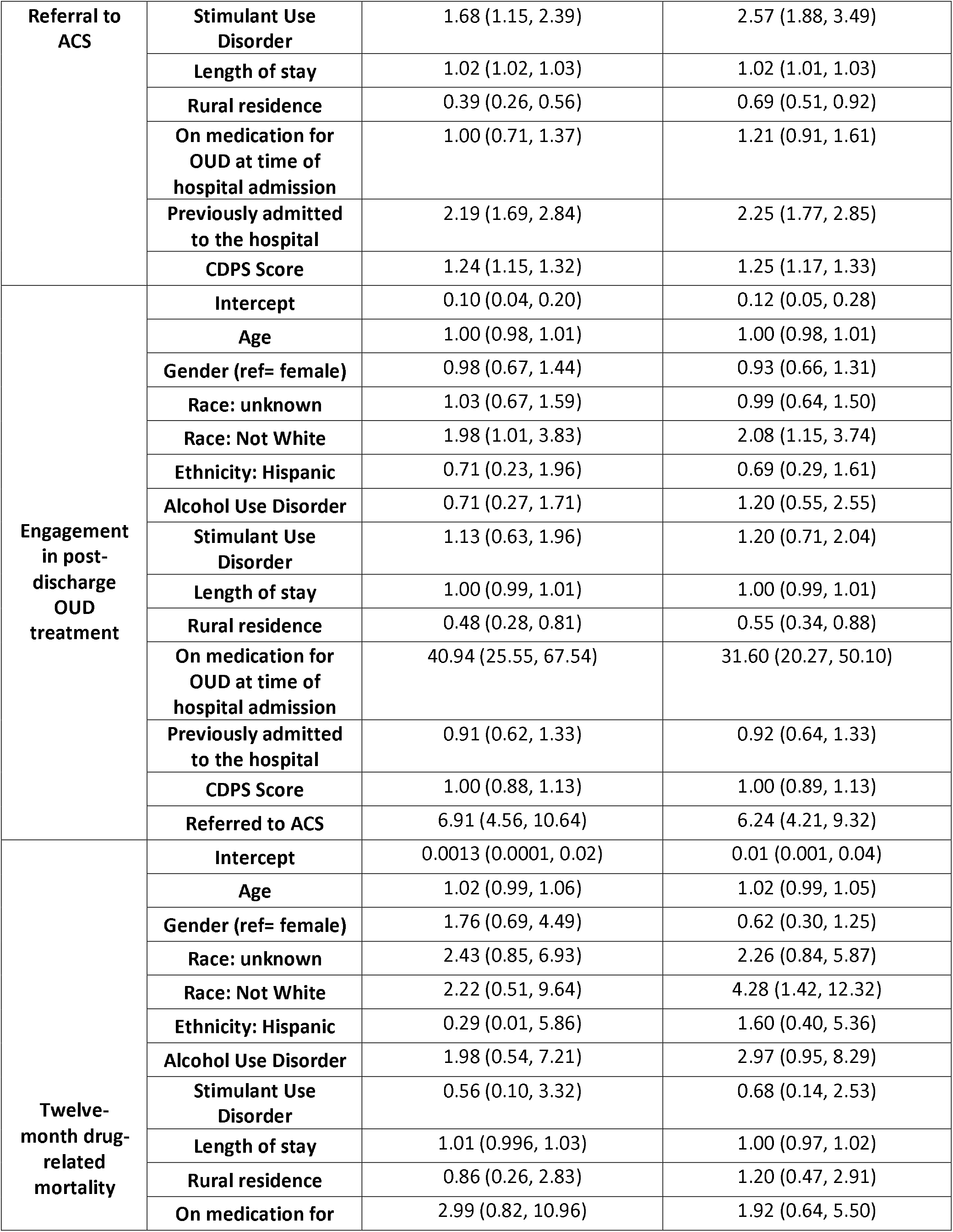
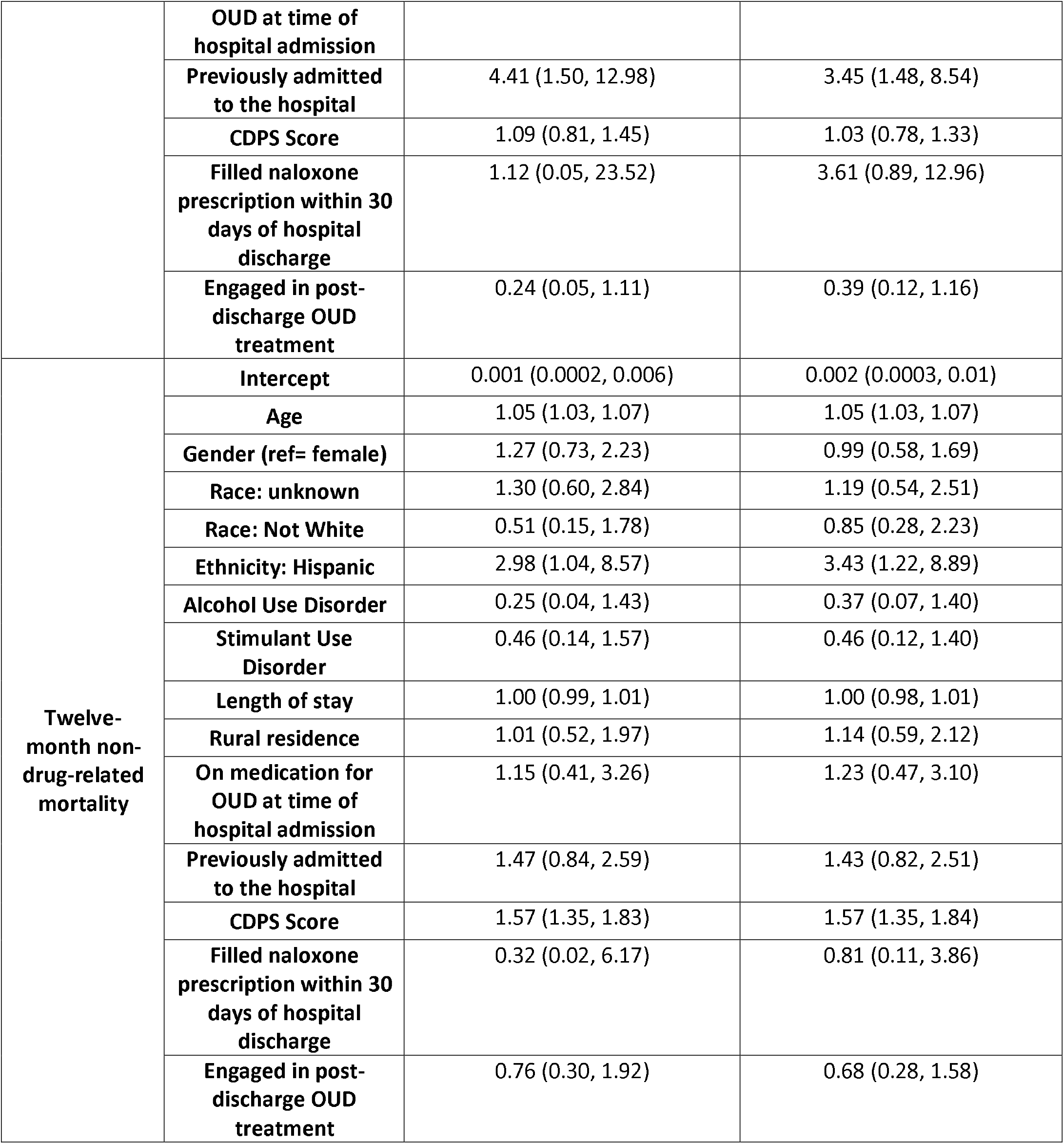

**Figure 1.**
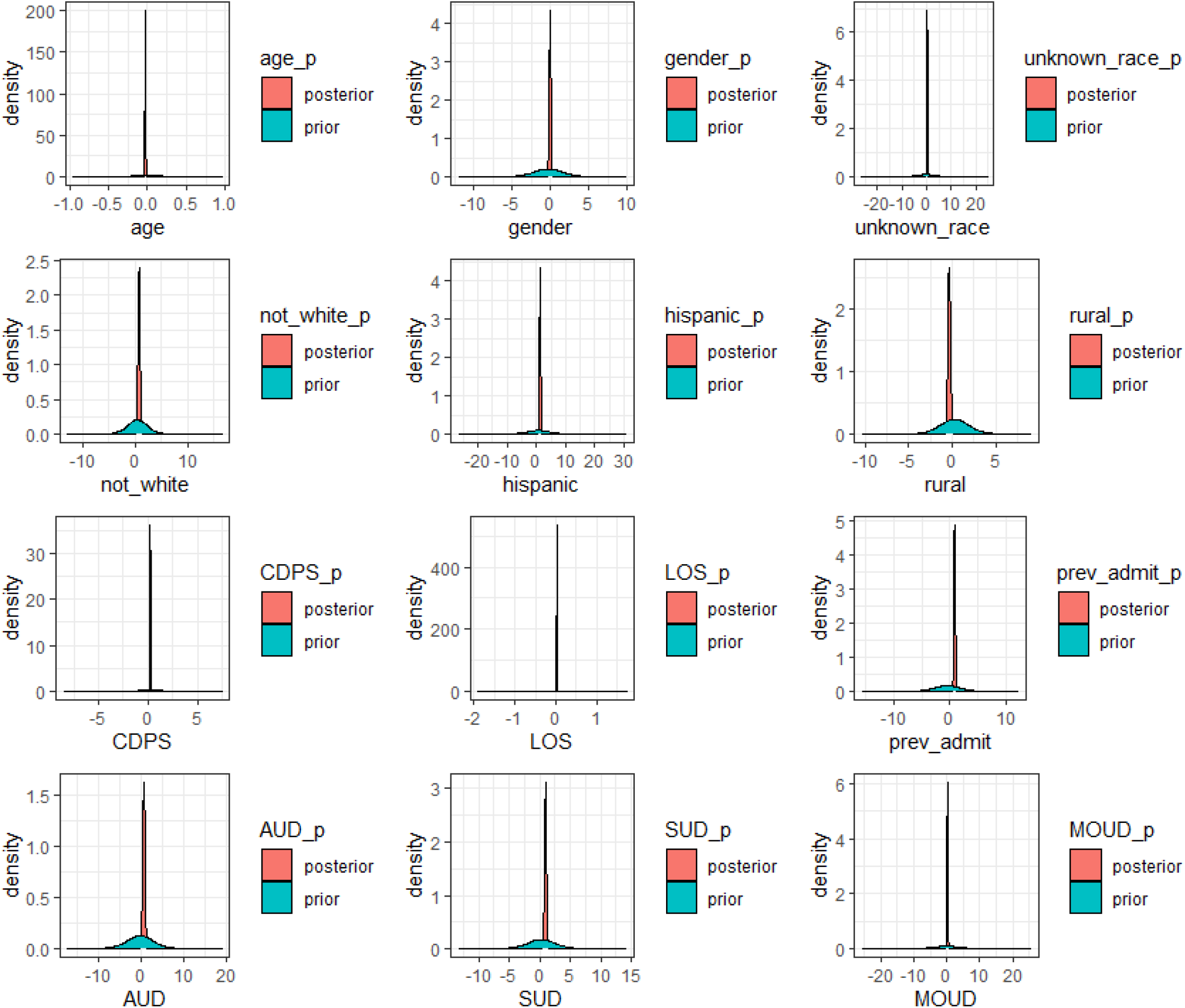
Prior-posterior plots for referral to addiction consult service.

**Figure 2.**
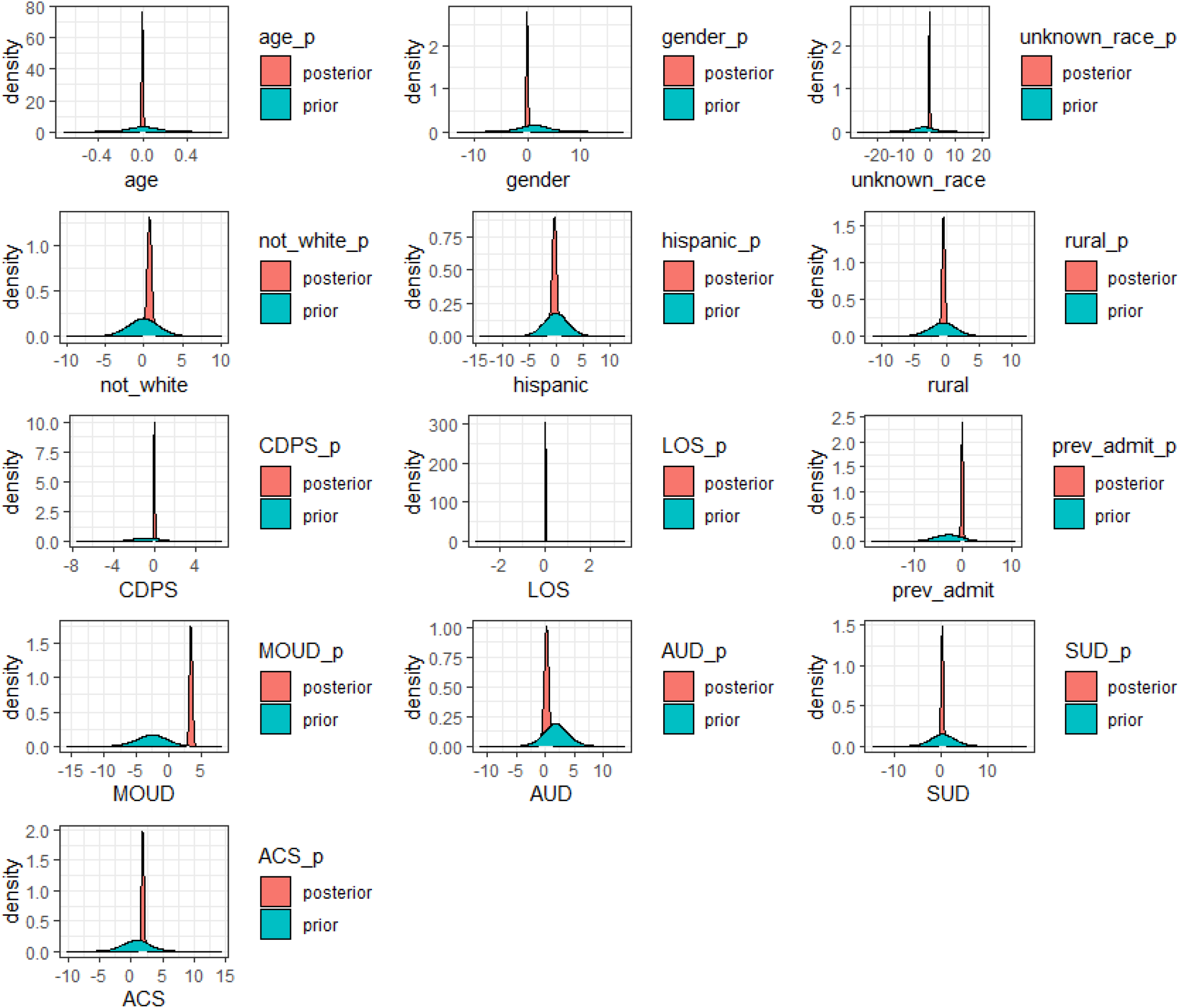
Prior-posterior plots for engagement in post-discharge OUD treatment.

**Figure 3.**
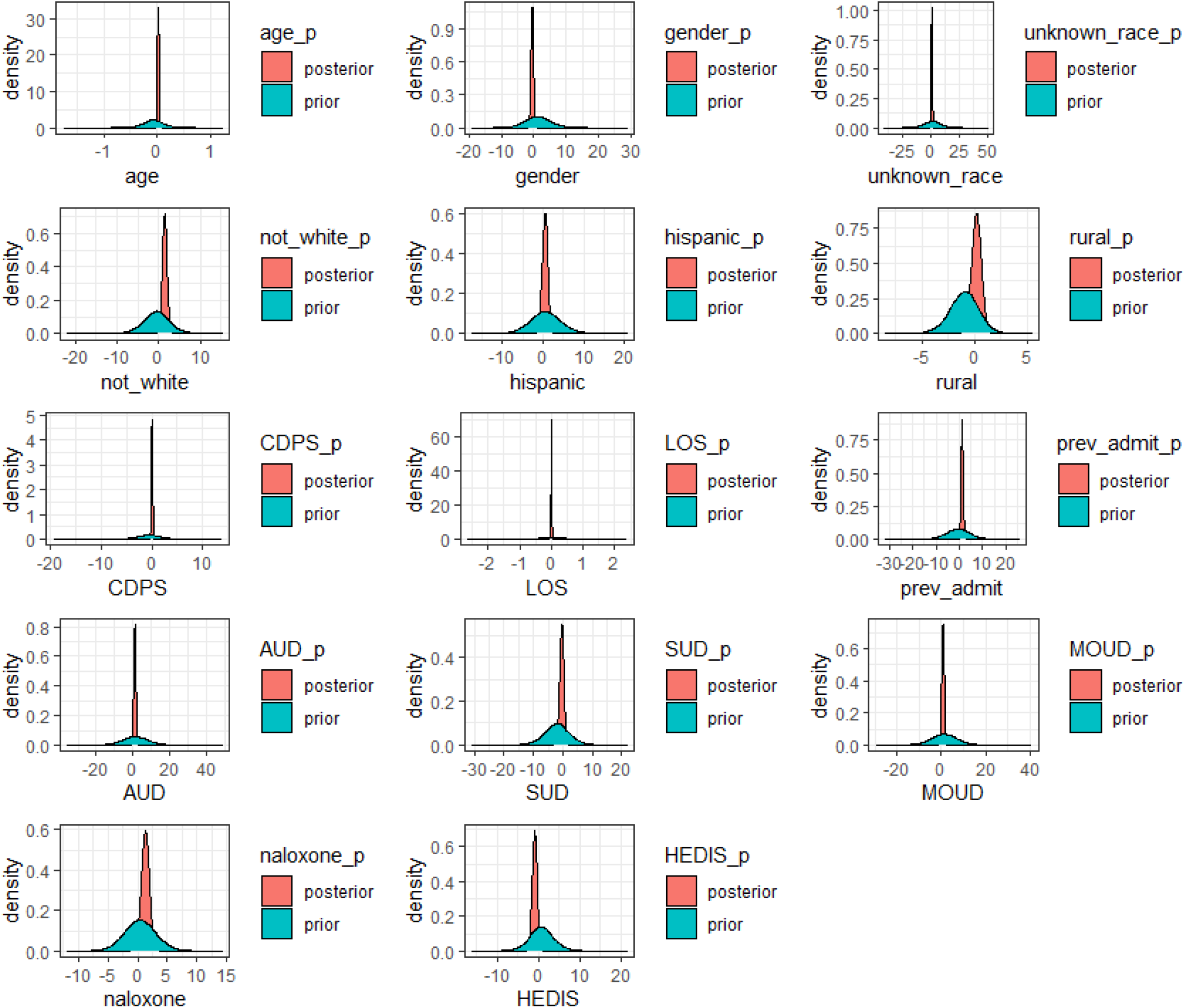
Prior-posterior plots for drug-related mortality at 12 months.

**Figure 4.**
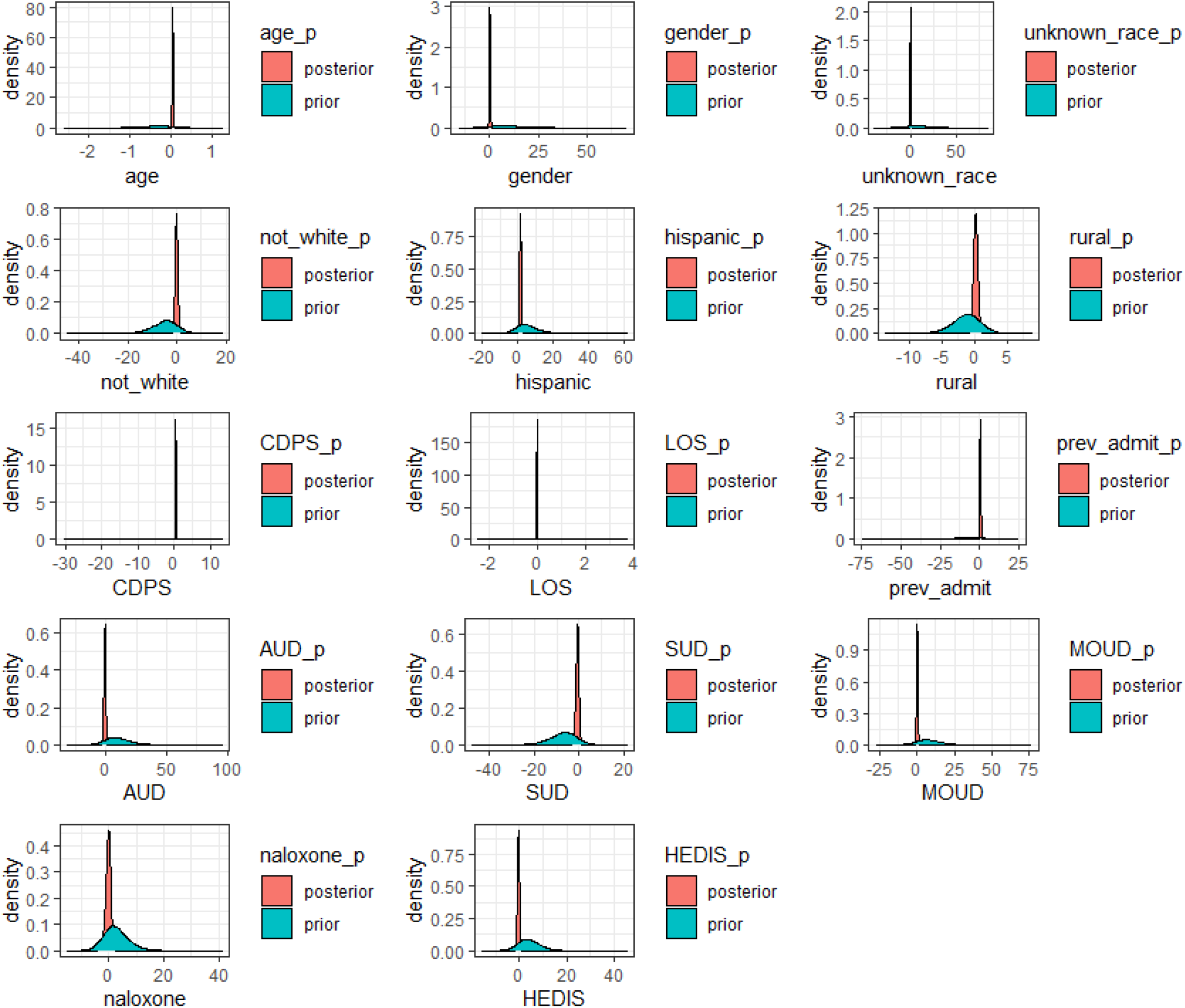
Prior-posterior plots for non-drug related mortality at 12 months.

### Appendix 3. Non-technical model description

#### Model and purpose

The purpose of this model is to understand how people who are hospitalized with opioid use disorder progress through care, from the time they are hospitalized through 12 months after they are discharged. We are especially interested in addiction care in Oregon, but the model could be used in states and settings other than ours.

#### Types of applications designed to address

The model is designed to understand mortality, from drug-related causes like overdose, and from non-drug related causes like heart attack, in the twelve-months after discharge from the hospital. We wanted to know how referral to addiction consult services, a specialized team in the hospital that cares for patients admitted with addiction, impact post-discharge engagement in treatment for opioid use disorder and death within twelve months of discharge.

#### Sources of funding and their role

This work was funded by the National Institutes of Health. The funder of the study had no role in study design, data collection, data analysis, data interpretation, or writing of the report. Dr. Korthuis serves as principal investigator for NIH-funded studies that accept donated study medication from Alkermes (extended-release naltrexone) and Indivior (buprenorphine).

#### Structure

Here is our model structure (Figure 1). Once patients were admitted to a hospital in Oregon from 2015 to 2018 and diagnosed with opioid use disorder, they could be referred to see an addiction consult service, or not. After they were discharged, they could engage in post-discharge care for opioid use disorder, or not. At twelve months, we looked to see if they were still alive, or if they had died, if it was from a drug-related, or non-drug related cause. We used Oregon Medicaid claims data to gather information. We also used information from experts in addiction. We combined the Medicaid data with the expert information using a technique called Bayesian analysis.

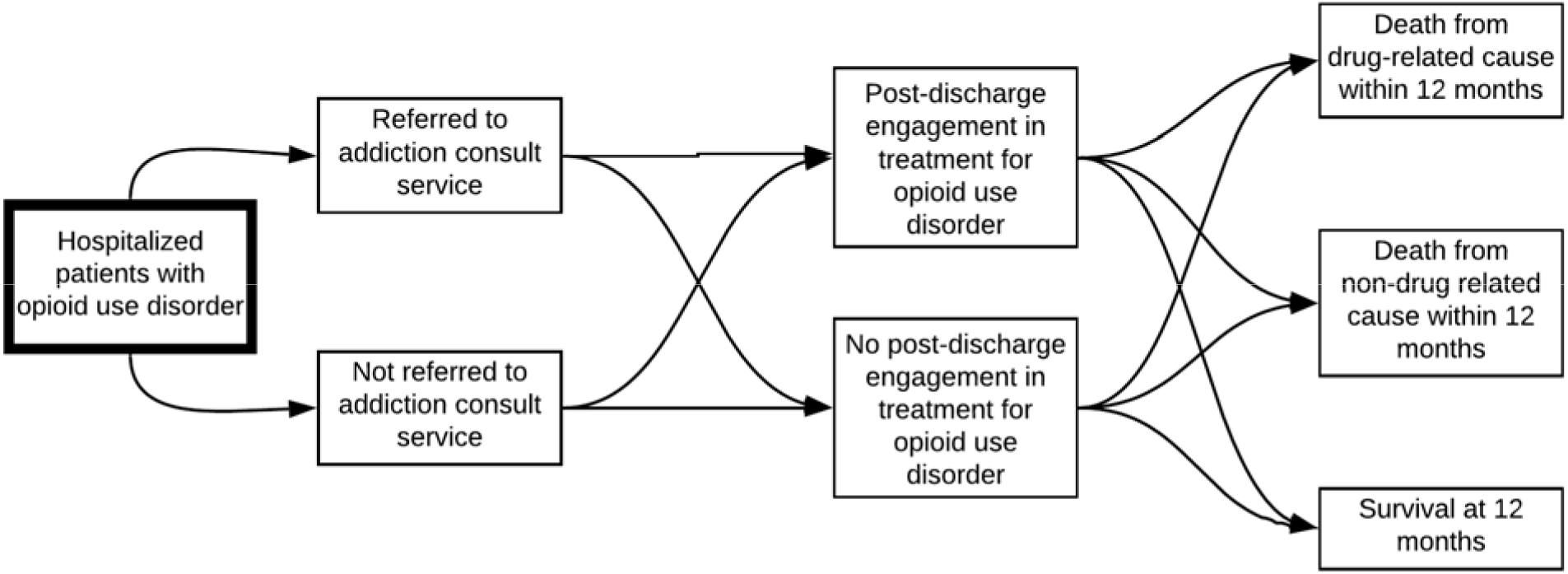

#### Model validation and summary of results

We validated our model a few ways. First, we spoke to experts about what we had planned, and they agreed that this model represented how patients move through care in real life, and that the questions we had about survival were important to answer. Next, once we fit our model, we checked to make sure our actual number of deaths matched the number modeled. Then, we compared our model estimates to another model, built independently by a research team in Boston, to compare results. After that, we used high-quality data from published studies to see if our model could accurately predict what happened to patients in published studies. We found that, in general, our model better matched observed estimates from Oregon than the national model, which suggests that using our model with local data in different contexts may provide more accurate information in those settings.

#### Main limitations for its intended applications

The main limitation of this model is that it does not include non-addiction consult service addiction care in hospitals. We are not able to tell how much of a difference there might be from addiction consult services versus standard addiction care provided by other types of doctors. However, we know that there are additional benefits from addiction consult services: they can help transform hospital environments more broadly to better care for patients with addiction. Additionally, very few people receive addiction care while in the hospital in general.

#### Reference to the model’s technical documentation

For more information, see our paper (cite).

## References

1. CDC NCHS. NCHS Data on Drug Poisoning Deaths. 2018.

2. Hser YI, Mooney LJ, Saxon AJ, Miotto K, Bell DS, Zhu Y, et al. High Mortality Among Patients With Opioid Use Disorder in a Large Healthcare System. J Addict Med. 2017;11(4):315–9. Epub 2017/04/21. doi: 10.1097/ADM.0000000000000312. PubMed PMID: 28426439; PubMed Central PMCID: PMCPMC5930020.

3. McNeil R, Small W, Wood E, Kerr T. Hospitals as a ‘risk environment’: an ethno-epidemiological study of voluntary and involuntary discharge from hospital against medical advice among people who inject drugs. Soc Sci Med. 2014;105:59–66. Epub 2014/02/11. doi: 10.1016/j.socscimed.2014.01.010. PubMed PMID: 24508718; PubMed Central PMCID: PMCPMC3951660.

4. Alfandre DJ. “I’m going home”: discharges against medical advice. Mayo Clin Proc. 2009;84(3):255–60. Epub 2009/03/03. doi: 10.1016/S0025-6196(11)61143-9. PubMed PMID: 19252113; PubMed Central PMCID: PMCPMC2664598.

5. Davoli M, Bargagli AM, Perucci CA, Schifano P, Belleudi V, Hickman M, et al. Risk of fatal overdose during and after specialist drug treatment: the VEdeTTE study, a national multi-site prospective cohort study. Addiction. 2007;102(12):1954–9. Epub 2007/11/23. doi: 10.1111/j.1360-0443.2007.02025.x. PubMed PMID: 18031430.

6. Noska A, Mohan A, Wakeman S, Rich J, Boutwell A. Managing Opioid Use Disorder During and After Acute Hospitalization: A Case-Based Review Clarifying Methadone Regulation for Acute Care Settings. J Addict Behav Ther Rehabil. 2015;4(2). Epub 2015/08/11. doi: 10.4172/2324-9005.1000138. PubMed PMID: 26258153; PubMed Central PMCID: PMCPMC4527170.

7. Wakeman SE, Larochelle MR, Ameli O, Chaisson CE, McPheeters JT, Crown WH, et al. Comparative Effectiveness of Different Treatment Pathways for Opioid Use Disorder. JAMA Netw Open. 2020;3(2):e1920622. Epub 2020/02/06. doi: 10.1001/jamanetworkopen.2019.20622. PubMed PMID: 32022884.

8. Priest KC, Lovejoy, T., Englander, H., Shull., S., McCarty, D. Opioid agonist therapy during hospitalization within the Veterans Health Administration: A retrospective cohort analysis.. JGIM. 2019. doi: Under review.

9. Jones CM, Campopiano M, Baldwin G, McCance-Katz E. National and State Treatment Need and Capacity for Opioid Agonist Medication-Assisted Treatment. Am J Public Health. 2015;105(8):e55–63. Epub 2015/06/13. doi: 10.2105/AJPH.2015.302664. PubMed PMID: 26066931; PubMed Central PMCID: PMCPMC4504312.

10. Englander H, King C, Nicolaidis C, Collins D, Patten A, Gregg J, et al. Predictors of Opioid and Alcohol Pharmacotherapy Initiation at Hospital Discharge Among Patients Seen by an Inpatient Addiction Consult Service. J Addict Med. 2019. Epub 2019/12/24. doi: 10.1097/ADM.0000000000000611. PubMed PMID: 31868830.

11. Englander H, Dobbertin K, Lind BK, Nicolaidis C, Graven P, Dorfman C, et al. Inpatient Addiction Medicine Consultation and Post-Hospital Substance Use Disorder Treatment Engagement: a Propensity-Matched Analysis. J Gen Intern Med. 2019. Epub 2019/08/15. doi: 10.1007/s11606-019-05251-9. PubMed PMID: 31410816.

12. Wakeman SE, Metlay JP, Chang Y, Herman GE, Rigotti NA. Inpatient Addiction Consultation for Hospitalized Patients Increases Post-Discharge Abstinence and Reduces Addiction Severity. J Gen Intern Med. 2017;32(8):909–16. Epub 2017/05/21. doi: 10.1007/s11606-017-4077-z. PubMed PMID: 28526932; PubMed Central PMCID: PMCPMC5515798.

13. Clark AK, Wilder CM, Winstanley EL. A systematic review of community opioid overdose prevention and naloxone distribution programs. J Addict Med. 2014;8(3):153–63. Epub 2014/05/31. doi: 10.1097/ADM.0000000000000034. PubMed PMID: 24874759.

14. Coffin PO, Sullivan SD. Cost-effectiveness of distributing naloxone to heroin users for lay overdose reversal. Ann Intern Med. 2013;158(1):1–9. Epub 2013/01/02. doi: 10.7326/0003-4819-158-1-201301010-00003. PubMed PMID: 23277895.

15. Irvine MA, Buxton JA, Otterstatter M, Balshaw R, Gustafson R, Tyndall M, et al. Distribution of take-home opioid antagonist kits during a synthetic opioid epidemic in British Columbia, Canada: a modelling study. Lancet Public Health. 2018;3(5):e218–e25. Epub 2018/04/22. doi: 10.1016/S2468-2667(18)30044-6. PubMed PMID: 29678561.

16. Irvine MA, Kuo M, Buxton JA, Balshaw R, Otterstatter M, Macdougall L, et al. Modelling the combined impact of interventions in averting deaths during a synthetic-opioid overdose epidemic. Addiction. 2019;114(9):1602–13. Epub 2019/06/06. doi: 10.1111/add.14664. PubMed PMID: 31166621; PubMed Central PMCID: PMCPMC6684858.

17. Weinstein ZM, Wakeman SE, Nolan S. Inpatient Addiction Consult Service: Expertise for Hospitalized Patients with Complex Addiction Problems. Med Clin North Am. 2018;102(4):587–601. Epub 2018/06/24. doi: 10.1016/j.mcna.2018.03.001. PubMed PMID: 29933817; PubMed Central PMCID: PMCPMC6750950.

18. McDuff DR, Solounias BL, Beuger M, Cohen A, Klecz M, Weintraub E. A substance abuse consultation service. Enhancing the care of hospitalized substance abusers and providing training in addiction psychiatry. Am J Addict. 1997;6(3):256–65. Epub 1997/07/01. doi: 10.3109/10550499709136993. PubMed PMID: 9256992.

19. Collins D, Alla J, Nicolaidis C, Gregg J, Gullickson DJ, Patten A, et al. “If It Wasn’t for Him, I Wouldn’t Have Talked to Them”: Qualitative Study of Addiction Peer Mentorship in the Hospital. Journal of general internal medicine. 2019. doi: 10.1007/s11606-019-05311-0. PubMed PMID: 31512181.

20. Englander H, Gregg J, Gullickson J, Cochran-Dumas O, Colasurdo C, Alla J, et al. Recommendations for integrating peer mentors in hospital-based addiction care. Subst Abus. 2019:1–6. Epub 2019/09/07. doi: 10.1080/08897077.2019.1635968. PubMed PMID: 31490736.

21. Englander H, Collins D, Perry SP, Rabinowitz M, Phoutrides E, Nicolaidis C. “We’ve Learned It’s a Medical Illness, Not a Moral Choice”: Qualitative Study of the Effects of a Multicomponent Addiction Intervention on Hospital Providers’ Attitudes and Experiences. J Hosp Med. 2018;13(11):752–8. doi: 10.12788/jhm.2993. PubMed PMID: 29694454.

22. Priest KC, McCarty D. Role of the Hospital in the 21st Century Opioid Overdose Epidemic: The Addiction Medicine Consult Service. J Addict Med. 2019;13(2):104–12. Epub 2019/01/05. doi: 10.1097/ADM.0000000000000496. PubMed PMID: 30608266; PubMed Central PMCID: PMCPMC6417955.

23. HEDIS. Initiation and Engagement of Alcohol and Other Drug Abuse or Dependence Treatment (IET) [cited 2020 October 20th]. Available from: https://www.ncqa.org/hedis/measures/initiation-and-engagement-of-alcohol-and-other-drug-abuse-or-dependence-treatment/.

24. McNeely J, Troxel AB, Kunins HV, Shelley D, Lee JD, Walley A, et al. Study protocol for a pragmatic trial of the Consult for Addiction Treatment and Care in Hospitals (CATCH) model for engaging patients in opioid use disorder treatment. Addict Sci Clin Pract. 2019;14(1):5. Epub 2019/02/20. doi: 10.1186/s13722-019-0135-7. PubMed PMID: 30777122; PubMed Central PMCID: PMCPMC6380041.

25. Englander H, Weimer M, Solotaroff R, Nicolaidis C, Chan B, Velez C, et al. Planning and Designing the Improving Addiction Care Team (IMPACT) for Hospitalized Adults with Substance Use Disorder. J Hosp Med. 2017;12(5):339–42. Epub 2017/05/02. doi: 10.12788/jhm.2736. PubMed PMID: 28459904; PubMed Central PMCID: PMCPMC5542562.

26. King C, Nicolaidis C, Korthuis PT, Priest KC, Englander H. Patterns of substance use before and after hospitalization among patients seen by an inpatient addiction consult service: A latent transition analysis. Journal of Substance Abuse Treatment. doi: 10.1016/j.jsat.2020.108121. PubMed PMID: 108121.

27. Bedrick EJ, Christensen R, Johnson W. Bayesian Binomial Regression: Predicting Survival at a Trauma Center. The American Statistician. 1997;51(3):211–8. doi: 10.1080/00031305.1997.10473965.

28. Hanmer MJ, Ozan Kalkan K. Behind the Curve: Clarifying the Best Approach to Calculating Predicted Probabilities and Marginal Effects from Limited Dependent Variable Models. American Journal of Political Science. 2013;57(1):263–77. doi: 10.1111/j.1540-5907.2012.00602.x.

29. Wheeler B. Package ‘AlgDesign’ 2011. Available from: https://cran.r-project.org/web/packages/AlgDesign/AlgDesign.pdf.

30. Carpenter B, Gelman A, Hoffman MD, Lee D, Goodrich B, Betancourt M, et al. Stan: A Probabilistic Programming Language. 2017. 2017;76(1):32. Epub 2017-01-11. doi: 10.18637/jss.v076.i01.

31. Stan Development Team. ShinyStan: Interactive Visual and Numerical Diagnostics and Posterior Analysis for Bayesian Models 2018. Available from: http://mc-stan.org.

32. Vehtari A, Gelman A, Gabry J. Practical Bayesian model evaluation using leave-one-out cross-validation and WAIC. Statistics and Computing. 2017;27(5):1413–32. doi: 10.1007/s11222-016-9696-4.

33. Scogin et al. BayesPostEst: An R Package to Generate Postestimation Quantities for Bayesian MCMC Estimation. Journal of Open Source Software. 2019;4(42):1722. doi: 10.21105/joss.01722.

34. Wickham H. ggplot2: Elegant Graphics for Data Analysis: Springer-Verlag New York; 2016.

35. Eddy DM, Hollingworth W, Caro JJ, Tsevat J, McDonald KM, Wong JB, et al. Model transparency and validation: a report of the ISPOR-SMDM Modeling Good Research Practices Task Force-7. Med Decis Making. 2012;32(5):733–43. Epub 2012/09/20. doi: 10.1177/0272989X12454579. PubMed PMID: 22990088.

36. Sendi PP, Craig BA, Pfluger D, Gafni A, Bucher HC. Systematic validation of disease models for pharmacoeconomic evaluations. Swiss HIV Cohort Study. J Eval Clin Pract. 1999;5(3):283–95. Epub 1999/08/26. doi: 10.1046/j.1365-2753.1999.00174.x. PubMed PMID: 10461580.

37. Barocas JA, Eftekhari Yazdi G, Savinkina A, Nolen S, Savitzky C, Samet JH, et al. Long-term infective endocarditis mortality associated with injection opioid use in the United States: a modeling study. Clin Infect Dis. 2020. Epub 2020/09/10. doi: 10.1093/cid/ciaa1346. PubMed PMID: 32901815.

38. Naeger S, Ali MM, Mutter R, Mark TL, Hughey L. Prescriptions Filled Following an Opioid-Related Hospitalization. Psychiatr Serv. 2016;67(11):1262–4. Epub 2016/11/02. doi: 10.1176/appi.ps.201500538. PubMed PMID: 27247179.

39. Larochelle MR, Bernson D, Land T, Stopka TJ, Wang N, Xuan Z, et al. Medication for Opioid Use Disorder After Nonfatal Opioid Overdose and Association With Mortality: A Cohort Study. Ann Intern Med. 2018;169(3):137–45. Epub 2018/06/19. doi: 10.7326/M17-3107. PubMed PMID: 29913516; PubMed Central PMCID: PMCPMC6387681.

40. Ashman J, DeFrances, C., Linman, S.,. Exploring hospital-based mortality – Examples from the 2014 National Hospital Care Survey Linked to the National Death Index: Distribution of in-hospital and post-acute mortality for patients hospitalized in 2014 2015. Available from: https://www.cdc.gov/nchs/data/nhcs/mortality_2014.pdf.

